# The Telesafe archive: creating a database of UK primary care telephone consultations

**DOI:** 10.64898/2026.05.19.26353559

**Authors:** Peter J Edwards, Barbara Caddick, Adam Skeen, Jordan Lin, Helena Thornton, Matthew J Ridd, Rebecca K Barnes, Chris Salisbury

**Affiliations:** Centre for Academic Primary Care, Bristol Medical School, University of Bristol; University of Birmingham; Somerset NHS Foundation Trust; Nuffield Department of Primary Care Health Sciences, University of Oxford

## Abstract

**Background:** In 2024, one-third of GP appointments in England were conducted by telephone. What happens during these consultations is largely unknown.

**Aim:** To test the feasibility of collecting recorded GP telephone consultations with linked data and consent for future research use.

**Design and setting:** Retrospective observational study in seven practices in South West England.

**Method:** Adults who had a telephone consultation at practices that routinely record calls were invited to consent to retrieval of call audio, a 4-month electronic health record (EHR) extract and a post-consultation patient questionnaire. Practice-level consent rates were analysed using regression models.

**Results:** Of 28 clinicians recruited, 19 GPs had consultations with patients whose recordings were retrievable, usable, and consented for future research. Of 2,053 invitations, 123 patients consented (6.0%). Consent was lower in more deprived practices (IMD 1–2 vs 9–10: OR=0.22, 95CI=0.09–0.54). Of 101 recordings retrieved, 96 were usable and 91 had consent for future research. 86/91 were linked to EHRs and 89/91 to post-consultation patient questionnaires. Mean consultation duration was 7 minutes 13 seconds; audible typing was heard in 69% (63/91). 161 problems were discussed (mean 1.77 per consultation). Most patients were happy their consultation was by telephone (96/117, 82%), although the majority reported usually preferring face-to-face appointments (68/115, 59%).

**Conclusion:** It is feasible to assemble a reusable archive of GP telephone consultations with linked data. However, recruitment was low using retrospective remote consent. Future work should test alternative recruitment approaches, particularly to improve patient engagement at practices serving deprived populations.

**How this fits in:**

- Telephone consultations now account for approximately one-third of GP appointments in England, yet the interactional detail of these calls remains under-described.
- This study establishes the One in a Million Telesafe archive: 91 recorded GP telephone consultations from seven practices, linked to EHR and patient-and clinician-reported data, with permissions for research re-use.
- Feasibility was demonstrated, but retrospective, remote consent produced low participation (6%) with lower responses in more deprived practices, limiting generalisability of estimates from this dataset.
- The archive offers a platform to examine communication quality and documentation, informing service design for safe, person-centred remote primary care.

## Introduction

In 2024, 367.4 million primary care appointments took place in England, averaging around one million per day.^1^ Despite this scale, the intricacies of these consultations remain relatively underexplored. Large-scale analyses of coded data offer statistical power to test associations but are limited by incomplete and potentially biased clinician coding.^2–4^ Qualitative interviews can provide in-depth individual accounts but are often shaped by a small number of participants’ perceptions and recall.^5^

The ‘One in a Million’ (OiaM) archive of video-recorded face-to-face GP consultations was collected in 2014–2015, its name underscoring the scale of daily primary care activity.^6,7^ The study included consultations with 327 unselected adult patients attending 23 GPs across 12 practices purposively sampled to capture variation in practice context. Recordings were linked to their medical record entries, pre-and post-consultation patient and GP surveys, and basic practice-level information. Although held as a controlled dataset, the OiaM archive has been very successful in its data sharing aim to support other research projects. To date it has been used in 20 subsequent studies generating 29 academic papers.

Since data collection for the OiaM archive, the use of telephone consultations has grown considerably. In 2014/15, around 14% of all primary care consultations were conducted by telephone;^7^ by 2024 this proportion had risen to 25%.^1^ When only GP appointments are considered, the rate is even higher – of 164.7 million GP appointments in 2024, one-third (54.3 million) were by telephone.^8,9^

Although telephone consultations were already increasing before the COVID-19 pandemic,^10^ this trend rapidly accelerated as practices adopted a ‘remote by default’ model. This raised safety concerns, and during the recovery stage there was widespread media criticism of the perceived overuse of telephone consultations.^11^ Today, rates are broadly similar to those seen immediately before the pandemic in 2019.^12^ Beyond rates, important differences exist between face-to-face and telephone GP consultations. Comparative analyses of recorded NHS GP consultations have shown that telephone consultations are typically shorter, address fewer problems, and involve less data gathering, counselling, and rapport-building than face-to-face consultations.^13,14^ Telephone consultations also lack visual cues, an important part of clinical assessment.^15^ These differences may alter clinician–patient interactions, underscoring the value of studying telephone consultations.

This project aimed (i) to explore the feasibility of creating a reusable dataset of recorded telephone consultations (the *One in a Million Telesafe* dataset); and (ii) to underpin a separate study of safety-netting advice in telephone consultations reported elsewhere.^16^ This data paper also describes the characteristics of the practices, patients, consultations and problems included in the final dataset.

## Methods

A cross-sectional observational design was employed, with retrospective collection of routinely recorded telephone consultations, combined with collection of routine data from medical records and questionnaires from participating clinicians and patients.

### Recruitment

Practices were recruited from the Bristol, North Somerset and South Gloucestershire (BNSSG) Integrated Care Board area via the NIHR Research Delivery Network. Eligibility criteria were: use of the EMIS electronic health record (EHR) system; routine audio recording of telephone consultations; and at least two clinicians who regularly conducted telephone consultations and consented to recordings being retained for the research dataset.

Patient recruitment took place in two waves. In the first wave (April–November 2023), five practices used a postal consent process, an approach recommended by a patient and public involvement (PPI) group of varied ages, genders, and ethnicities. Each week practices ran an EMIS search to identify all patients aged 18 or over who had a telephone consultation with a participating clinician in the preceding week. Clinicians then screened the list against predefined exclusion criteria: patients receiving end-of-life care, consultations on behalf of a third party, patients unable to consult in English, and those the clinician deemed inappropriate to invite. As screening was weekly and invitations were posted, patients typically received them 1–3 weeks after their consultation.

Because of low recruitment, an alternative strategy was introduced for the second wave (November 2023–June 2024) using an electronic process. To notify patients more quickly and align with usual practice communication, clinicians or administrators sent eligible patients a text message shortly after their consultation. The message linked to the study website where patients could read the participant information leaflet and complete an electronic consent form, with the option to request paper copies.

### Surveys

Clinicians completed a 15-question survey at enrolment: 10 items on demographics and professional background and 5 on safety-netting (Supplementary materials). Patients completed a post-consultation survey with 20 questions covering demographics; preferences and expectations; experience and satisfaction with the index consultation; and safety-netting recall (Supplementary materials).

### Medical records

EHR data were extracted for the index consultation and all appointments in the period 1 month before to 3 months after this, together with a summary of long-term conditions.

### Consent

Participants gave separate consent for retention of each data type for further research (audio recording, transcript, survey responses and medical records), and separately for use in teaching.

### Consultation contents

Consultation durations were exported to the nearest second for each recording, then manually checked. Timings were adjusted to exclude phone dialling and any period when another person answered before handing the phone to the patient.

Problems were coded from the consultation recordings using the Complex Consultations Coding Toolkit^17^ and the International Classification of Primary Care, version 3 (ICPC-3).^18,19^ Coding was undertaken by AS, CS, JL and HT, and double-checked by the first author (PE).

### Software

Research Electronic Data Capture (REDCap) was used to administer electronic questionnaires. Statistical analysis was conducted in Stata/MP 18.0, with code available online.^9^ Mixed-effects logistic regression (random intercept for practice) modelled consents out of invitations by practice and invitation mode, with IMD quintile, invitation mode and the practice-level percentage of patients of non-white ethnicity as predictors. Odds ratios (ORs) with 95% confidence intervals (CIs) are reported.

## Results

Figure 1 shows the participant recruitment flow. Table 1 summarises practice, clinician and patient demographics in the archive.

**Figure 1.**
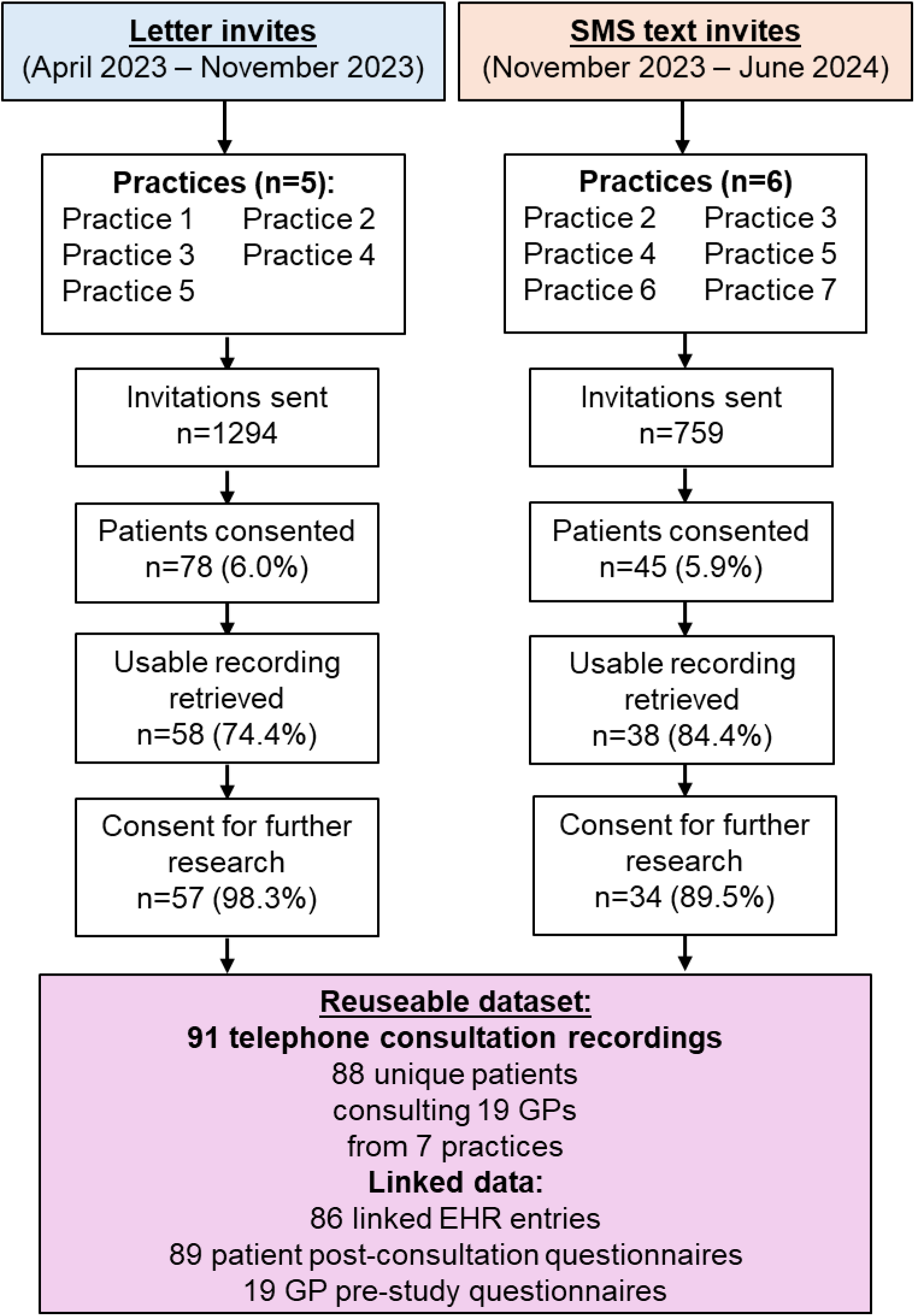
Recruitment flow into the reusable dataset. EHR = Electronic Health Records. GP = General Practitioner

**Table 1.** Practice, GP, patient and consultation characteristics. N=91 consultations with 88 unique patients (*excludes duplicates of the 3 patients who have 2 consultations in the archive).

| **Practice IMD decile (n=7)** | **n** | **%** |  | **Consultations per practice IMD decile (n=91)** | **n** | **%** |
| --- | --- | --- | --- | --- | --- | --- |
| 1 (most deprived) | 2 | 29% |  | 1 | 19 | 21% |
| 2 | 1 | 14% |  | 2 | 7 | 8% |
| 5 | 2 | 29% |  | 5 | 6 | 7% |
| 9 | 1 | 14% |  | 9 | 42 | 46% |
| 10 (least deprived) | 1 | 14% |  | 10 | 17 | 19% |
| **Practice list size (n=7)** |  |  |  | **No of problems per consultation (n=91)** |  |  |
| 5 to ≤10k | 1 | 14% |  | 1 | 48 | 53% |
| 10 to ≤15k | 2 | 29% |  | 2 | 22 | 24% |
| 15 to ≤20k | 1 | 14% |  | 3 | 17 | 19% |
| 20 to ≤30k | 1 | 14% |  | ≥4 | 4 | 4% |
| 30 to ≤40k | 2 | 29% |  |  |  |  |
| **Practice population non-white ethnicity (n=7)** |  |  |  | **Consultations with (n=91)** |  |  |
| <5% | 3 | 43% |  | GP Partner | 83 | 91% |
| 5 to ≤10% | 1 | 14% |  | Salaried GP | 7 | 8% |
| 10 to ≤15% | 3 | 43% |  | GP registrar | 1 | 1% |
| **GP gender (n=19)** |  |  |  | **Patient gender (n=88)\*** |  |  |
| Male | 8 | 42% |  | Male | 33 | 38% |
| Female | 11 | 58% |  | Female | 55 | 63% |
| **GP age (n=19)** |  |  |  | **Patient age (n=88)\*** |  |  |
| 18-34 | 2 | 11% |  | 18-34 | 8 | 9% |
| 35-49 | 12 | 63% |  | 35-49 | 9 | 10% |
| 50-64 | 5 | 26% |  | 50-64 | 21 | 24% |
| >65 | 0 | 0% |  | >65 | 49 | 56% |
| Not reported | 0 | 0% |  | Not reported | 1 | 1% |
| **GP ethnicity (n=19)** |  |  |  | **Patient ethnicity (n=88)\*** |  |  |
| White | 17 | 89% |  | White | 81 | 92% |
| Other | 2 | 11% |  | Other | 2 | 2% |
| Not reported / prefer not to say | 0 | 0% |  | Not reported / prefer not to say | 6 | 7% |

### Practice recruitment

Five practices were originally recruited and participated during the letter invites stage. One practice withdrew and was replaced by 2 additional practices in the text invite stage (Figure 1).

### Clinician recruitment

Twenty-eight clinicians consented to participate (21 GPs, 1 GP registrar, 4 nurse practitioners, 1 paramedic and 1 first contact physiotherapist). Of these, 19 clinicians (18 GPs and 1 GP registrar) had at least one retrievable consultation recording from a patient who consented to archiving for future research. Unless stated otherwise, results refer only to archived data. The mean clinician age was 45 years (SD=10.3, range=32-62), most were of White ethnicity (17/19) and GP Partners (14, Salaried GPs 4, GP registrar 1).

### Patient recruitment

A total of 2,053 invitations (1294 letters, 759 texts) were sent; 123 (6.0%) consent forms were returned – 78/1294 (6.0%) from letter invites and 45/759 (5.9%) from text invites. Of those who consented, 112/123 (91.1%) permitted use of their consultation recording, EHR and questionnaire for future research. Recordings were retrieved for 101 consultations; 5 were unusable (1 answerphone, 1 cut out mid-consultation, 2 with substantial third-party contributions, 1 arranged call-back with no additional recording available). Of the 96 usable recordings, 91 included consent for future research. Three patients contributed two consultations, yielding 91 consultations from 88 unique patients. The mean patient was 63.5 years (SD=16.1, range=18-94), most were female (63.4%, 55/88) and White ethnicity (92.0%, 81/88). Linked data were available for 83/91 consultations EHR entries (86 EHRs were retrieved, but 3 had no matching consultation entry on the index date) and for 89/91 post-consultation questionnaires

Overall, consent at the practice-level ranged from 2.1–11.8%. The highest rate observed in any round was 14.7% (letter round at one practice) and the lowest was 1.4% (text round at another; Table 2). Compared with the least deprived practices (IMD deciles 9 – 10, reference), the most deprived (IMD deciles 1–2) had lower odds of returning a consent form (OR=0.22, CI=0.09–0.54, p=0.001), as did middle-deprivation practices (IMD decile 5: OR=0.18, CI=0.08–0.42).

**Table 2.** Practice consent rates. *Practice non-white population shown to nearest 0.5% to protect identity of practices. N = 123 patients consent forms returned. Pract. = Practice. HCP = Health Care Professional.

| Prac. ID | Prac. IMD decile | HCPs consents | HCPs with recording | Letter invites | Letter consents |  | Text invites | Text consents |  | Overall consent rate |
| --- | --- | --- | --- | --- | --- | --- | --- | --- | --- | --- |
|  |  | (n) | (n) | (n) | (n) | (%) | (n) | (n) | (%) | (%) |
| 1 | 1 | 3 | 2 | 280 | 6 | 2.1% | 0 | - | - | 2.1% |
| 2 | 5 | 4 | 4 | 300 | 12 | 4.0% | 74 | 1 | 1.4% | 3.5% |
| 3 | 9 | 4 | 4 | 300 | 44 | 14.7% | 165 | 11 | 6.7% | 11.8% |
| 4 | 2 | 7 | 2 | 210 | 7 | 3.3% | 92 | 2 | 2.2% | 3.0% |
| 5 | 1 | 3 | 1 | 204 | 9 | 4.4% | 142 | 8 | 5.6% | 4.9% |
| 6 | 5 | 2 | 2 | 0 | - | - | 100 | 1 | 1.0% | 1.0% |
| 7 | 10 | 5 | 4 | 0 | - | - | 186 | 22 | 11.8% | 11.8% |

### Consultation characteristics

The mean consultation duration was 7 minutes:13 seconds (SD 4:14; range 1:01–24:14). Consultations where patients reported having their telephone appointment the same day they requested it were shorter (mean 5:50, SD 4:09, range 1:01–24:14) than those where patients waited at least one day (mean 8:16, SD 4:19, range 1:23–17:48).

Audible typing was present in 69.2% (63/91) of consultations. In most cases (58/63), typing occurred while the GP documented information gathered from the patient. In a few cases, typing appeared to be limited to entering prescription details (3/63), booking a follow-up appointment (1/63) or looking up information (1/63).

Across 91 consultations, 161 problems were discussed (mean 1.77 per consultation, SD 0.98; median 1; range 1–5). Table 3 summarises the types of problems discussed. The four most common ICPC-3 categories were *General* 19.3% (31/161) of all problems; 29.7% (27/91) of consultations, *Digestive* 10.6% (17/161); 15.4% (14/91), *Genital* 10.6% (17/161); 17.6% (16/91) and *Musculoskeletal* 10.6% (17/161); 17.6% (16/91).

**Table 3.** ICPC-3 Problem categories. *% of consultations in archive accounts for if there are 2 of the same problem categories in the same consultation. ICPC-3 = International Classification of Primary Care Version 3.

| Problem Type (ICPC-3) | Total in all recorded consultations | % of problems | Number of consultations with this problem type |
| --- | --- | --- | --- |
| **A - General** | 31 | 19.3% | 27 |
| **B - Blood, blood-forming organs and immune system** | 3 | 1.9% | 3 |
| **D - Digestive system** | 17 | 10.6% | 14 |
| **F - Eye** | 2 | 1.2% | 2 |
| **G - Genital system** | 17 | 10.6% | 16 |
| **H - Ear** | 2 | 1.2% | 2 |
| **K - Circulatory system** | 16 | 9.9% | 16 |
| **L - Musculoskeletal system** | 17 | 10.6% | 16 |
| **N - Neurological system** | 6 | 3.7% | 6 |
| **P - Psychological, mental and neurodevelopmental** | 7 | 4.3% | 7 |
| **R - Respiratory system** | 6 | 3.7% | 6 |
| **S - Skin** | 12 | 7.5% | 11 |
| **T - Endocrine, metabolic and nutritional system** | 13 | 8.1% | 13 |
| **U - Urinary system** | 9 | 5.6% | 9 |
| **W - Pregnancy and childbearing** | 2 | 1.2% | 2 |
| **Z - Social problems** | 1 | 0.6% | 1 |
| **Total** | 161 |  |  |

### Survey data

Table 4 summarises patient questionnaire results post-consultation for all respondents (n=120) and those included in the archive (n=89). Unless otherwise stated, results refer to all respondents. Responses from archive participants can be used alongside linked recordings and medical records to explore which consultation features are associated with specific patient experiences.

**Table 4.**
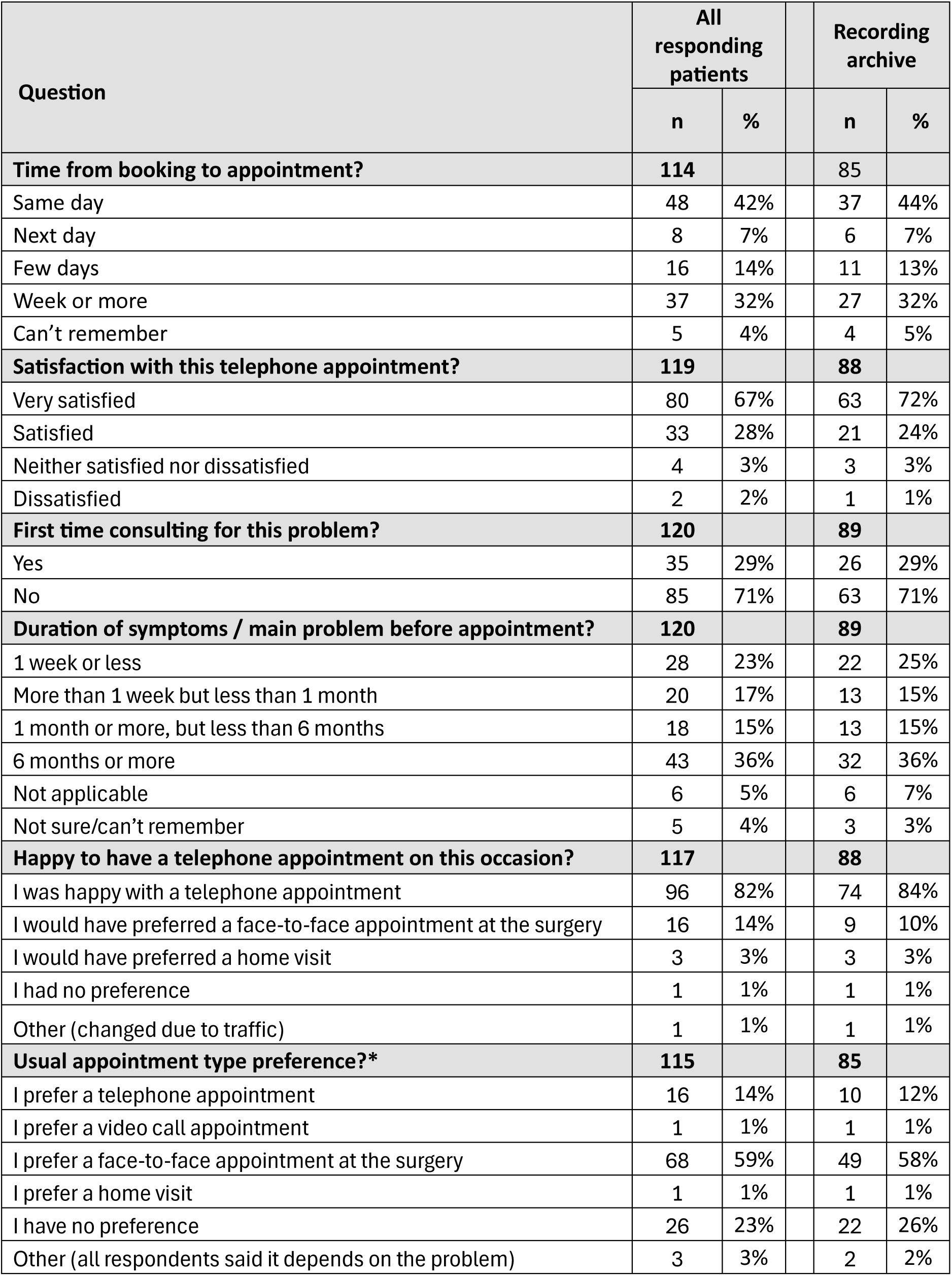
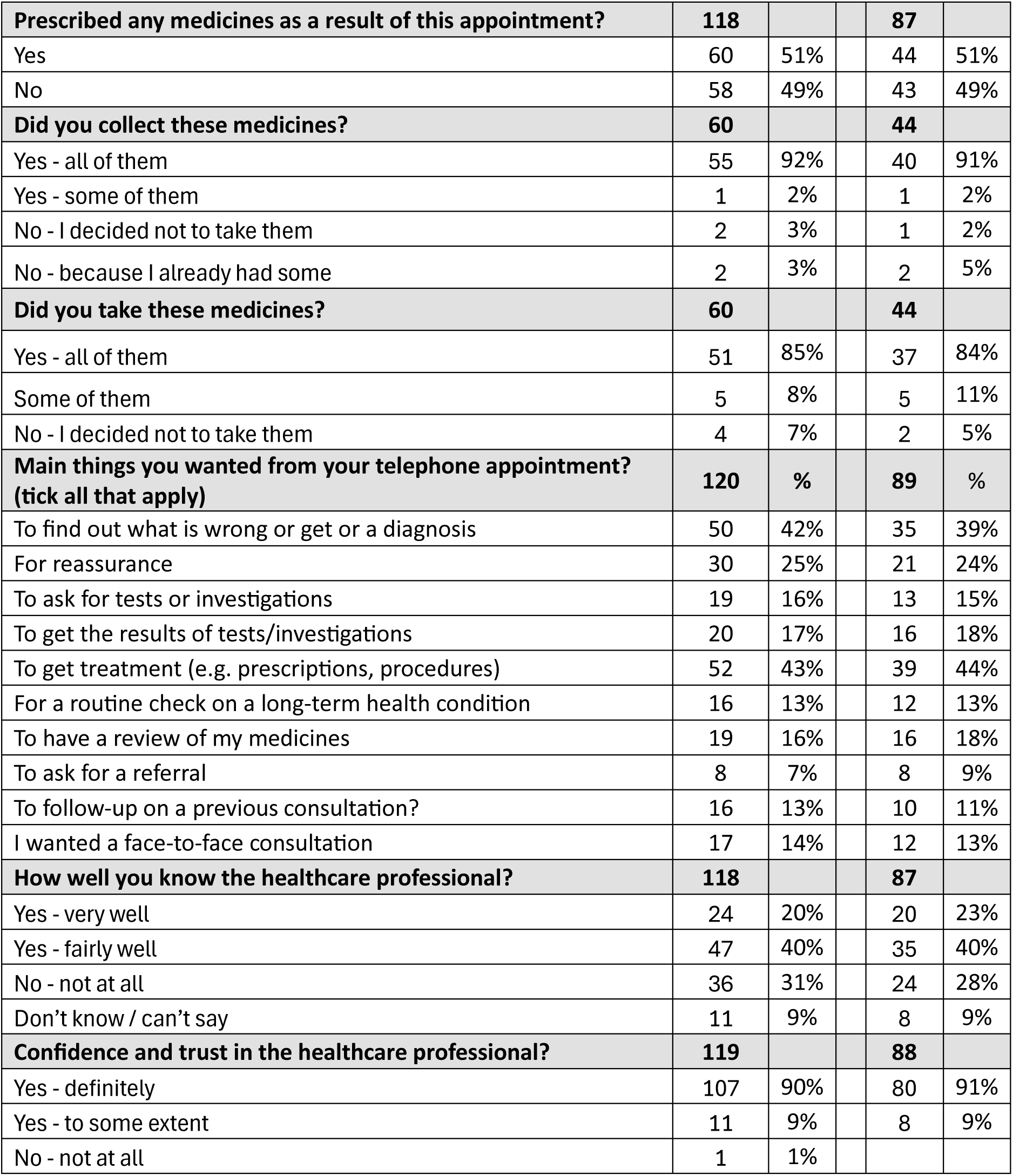
Patient survey results. *Excludes the second survey from the 3 patients with two consultations in the recording archive.

| Question | All responding patients |  | Recording archive |  |
| --- | --- | --- | --- | --- |
|  | n | % | n | % |
| **Time from booking to appointment?** | **114** |  | **85** |  |
| Same day | 48 | 42% | 37 | 44% |
| Next day | 8 | 7% | 6 | 7% |
| Few days | 16 | 14% | 11 | 13% |
| Week or more | 37 | 32% | 27 | 32% |
| Can't remember | 5 | 4% | 4 | 5% |
| **Satisfaction with this telephone appointment?** | **119** |  | **88** |  |
| Very satisfied | 80 | 67% | 63 | 72% |
| Satisfied | 33 | 28% | 21 | 24% |
| Neither satisfied nor dissatisfied | 4 | 3% | 3 | 3% |
| Dissatisfied | 2 | 2% | 1 | 1% |
| **First time consulting for this problem?** | **120** |  | **89** |  |
| Yes | 35 | 29% | 26 | 29% |
| No | 85 | 71% | 63 | 71% |
| **Duration of symptoms / main problem before appointment?** | **120** |  | **89** |  |
| 1 week or less | 28 | 23% | 22 | 25% |
| More than 1 week but less than 1 month | 20 | 17% | 13 | 15% |
| 1 month or more, but less than 6 months | 18 | 15% | 13 | 15% |
| 6 months or more | 43 | 36% | 32 | 36% |
| Not applicable | 6 | 5% | 6 | 7% |
| Not sure/can't remember | 5 | 4% | 3 | 3% |
| **Happy to have a telephone appointment on this occasion?** | **117** |  | **88** |  |
| I was happy with a telephone appointment | 96 | 82% | 74 | 84% |
| I would have preferred a face-to-face appointment at the surgery | 16 | 14% | 9 | 10% |
| I would have preferred a home visit | 3 | 3% | 3 | 3% |
| I had no preference | 1 | 1% | 1 | 1% |
| Other (changed due to traffic) | 1 | 1% | 1 | 1% |
| **Usual appointment type preference?\*** | **115** |  | **85** |  |
| I prefer a telephone appointment | 16 | 14% | 10 | 12% |
| I prefer a video call appointment | 1 | 1% | 1 | 1% |
| I prefer a face-to-face appointment at the surgery | 68 | 59% | 49 | 58% |
| I prefer a home visit | 1 | 1% | 1 | 1% |
| I have no preference | 26 | 23% | 22 | 26% |
| Other (all respondents said it depends on the problem) | 3 | 3% | 2 | 2% |

|  |  |  |  |  |  |
| --- | --- | --- | --- | --- | --- |
| **Prescribed any medicines as a result of this appointment?** | **118** |  |  | **87** |  |
| Yes | 60 | 51% |  | 44 | 51% |
| No | 58 | 49% |  | 43 | 49% |
| **Did you collect these medicines?** | **60** |  |  | **44** |  |
| Yes - all of them | 55 | 92% |  | 40 | 91% |
| Yes - some of them | 1 | 2% |  | 1 | 2% |
| No - I decided not to take them | 2 | 3% |  | 1 | 2% |
| No - because I already had some | 2 | 3% |  | 2 | 5% |
| **Did you take these medicines?** | **60** |  |  | **44** |  |
| Yes - all of them | 51 | 85% |  | 37 | 84% |
| Some of them | 5 | 8% |  | 5 | 11% |
| No - I decided not to take them | 4 | 7% |  | 2 | 5% |
| **Main things you wanted from your telephone appointment?**<br>**(tick all that apply)** | **120** | **%** |  | **89** | **%** |
| To find out what is wrong or get or a diagnosis | 50 | 42% |  | 35 | 39% |
| For reassurance | 30 | 25% |  | 21 | 24% |
| To ask for tests or investigations | 19 | 16% |  | 13 | 15% |
| To get the results of tests/investigations | 20 | 17% |  | 16 | 18% |
| To get treatment (e.g. prescriptions, procedures) | 52 | 43% |  | 39 | 44% |
| For a routine check on a long-term health condition | 16 | 13% |  | 12 | 13% |
| To have a review of my medicines | 19 | 16% |  | 16 | 18% |
| To ask for a referral | 8 | 7% |  | 8 | 9% |
| To follow-up on a previous consultation? | 16 | 13% |  | 10 | 11% |
| I wanted a face-to-face consultation | 17 | 14% |  | 12 | 13% |
| **How well you know the healthcare professional?** | **118** |  |  | **87** |  |
| Yes - very well | 24 | 20% |  | 20 | 23% |
| Yes - fairly well | 47 | 40% |  | 35 | 40% |
| No - not at all | 36 | 31% |  | 24 | 28% |
| Don't know / can't say | 11 | 9% |  | 8 | 9% |
| **Confidence and trust in the healthcare professional?** | **119** |  |  | **88** |  |
| Yes - definitely | 107 | 90% |  | 80 | 91% |
| Yes - to some extent | 11 | 9% |  | 8 | 9% |
| No - not at all | 1 | 1% |  |  |  |

About half of patients (49.1%, 56/114) reported consulting on the same or next day after making a request. Satisfaction with the index telephone appointment was high: 67% (80/119) were very satisfied and 28% (33/119) satisfied. Most (71%, 85/120) reported the index consultation was not their first for the problem. Around half (50.8%, 61/120) had had symptoms for a month or more. Acceptability of the telephone modality was high: 82% (96/117) were happy to consult by telephone, while 14% (16/117) would have preferred face-to-face. In general, however, most (59%, 68/115) usually preferred face-to-face consultations, with fewer preferring telephone (14%, 16/115) or having no preference (23%, 26/115).

Patients reported that medicines were prescribed in 51% (60/118) of consultations, but 8% (5/60) did not collect all items prescribed and 15% (9/60) did not take all of them. The most common patient goals were to obtain a diagnosis (42%, 50/120) or treatment (43%, 52/120). Most (60.1%, 71/118) reported knowing the healthcare professional very well or fairly well, though 31% (36/118) did not know them at all. The vast majority (90%, 107/119) reported ‘*definitely’* having confidence and trust in the healthcare professional.

## Discussion

### Summary

An archive of 91 audio-recorded GP telephone consultations from seven practices (19 GPs) has been established, covering 161 problems and linked to patient questionnaires and EHRs. Consultations averaged just over 7 minutes; about half addressed a single problem and half multiple problems. Nearly half occurred on the day patients requested care. Keyboard typing was audible in two-thirds of consultations, usually while patients provided information. Most patients were satisfied with their consultation and happy with a telephone appointment on this occasion, though their usual preference was face-to-face. Recruitment of practices and clinicians was feasible, but patient consent via retrospective remote contact was modest (6% overall) and lower in more deprived practices. Despite these limitations, the dataset spans a wide range of patients and clinical problems, offering potential to inform research on telephone consultations.

### Strengths and limitations

Although there have been previous studies of the content of telephone consultations in primary care,^20,21^ and surveys of patients’ experiences,^22^ the unique strength of this study is the availability of linked data from recorded consultations, medical records and patient questionnaires, with consent for re-use in future research. This allows examination of, for example, the relationship between how consultations are conducted and patient experience, or between consultation content and medical record documentation. This paper describes the range of data available for future studies.

Problem types were coded using established schemes by two authors. Although clinicians knew consultations might be included, they were unsure which patients would ultimately participate, and recruitment occurred over a long period, potentially reducing Hawthorne effects.^23^

The main limitation concerns recruitment, with practice participation limited to one area and most participating clinicians being GP partners. Although six non-GP professionals were recruited, no patients consulting with them consented to join the study. Patient participation required literacy, consulting in English, and post-event consent. Consent rates were lower in more deprived practices, and overall return of consent forms was poor, increasing the risk of responder bias and limiting generalisability. No information was collected on invitees who declined. Clinicians could also exclude patients deemed unsuitable for any reason, which may have introduced bias if those with unsatisfactory consultations were excluded.

### Comparison with existing literature

Average consultation duration in this dataset was longer than in audio-recorded Scottish GP telephone consultations from 2009 (4.6 minutes)^20^ and follow-up telephone consultations from 2017–18 (5.56 minutes).^21^ It was also longer than a large-scale analysis of English GP telephone consultations in 2013–14 (5.32 minutes),^24^ but shorter than out-of-hours NHS GP consultations collected in 2011 (7.78 minutes)^25^. Differences may partly reflect secular increases in consultation length over time and the later data collection in this study.^26^ Although telephone consultations may be shorter than face-to-face encounters, any efficiency gains may be offset by higher re-consultation rates, reduced opportunistic health promotion and concerns about the risk of patient harm if used inappropriately.^27,28^

Most patients in this study were satisfied or very satisfied with their appointment, consistent with findings among consenting participants in the OiaM study.^29^ However, ‘satisfied’ and ‘very satisfied’ are not interchangeable: patients often use ‘satisfied’ for care that was sufficient rather than outstanding, whereas ‘very satisfied’ denotes better-than-average to outstanding care allowing for meaningful comparisons.^30^ Conversely, consent among patients invited was much lower in this study (6%) compared to the OiaM study (79%), where a researcher was available on site to discuss the study face-to-face.

The frequency of problem types differed in this dataset compared with the OiaM study. Here, the most common problem type was ‘General’ (which includes polypharmacy reviews, abnormal results, cardiovascular risk assessments), whereas in the OiaM study it was musculoskeletal. Respiratory and psychological problems were also less frequent in this dataset.

### Implications for research and practice

This study has generated a controlled dataset that can be reused by other researchers, subject to ethical approvals, to better understand communication in telephone consultations. Feasibility has been demonstrated in South West England, and value could be enhanced by increasing participant diversity and replicating collection in other regions. Recruitment could be improved through prospective enrolment and researcher-supported contact by telephone rather than text or letter alone. Targeted approaches are also needed to address engagement inequities, for example through community outreach and collaboration with Deep End networks.^31^

The sound of typing was audible in more than two-thirds of consultations. Patients commonly dislike computer use during clinical encounters.^32^ When these consultations were recorded, ambient AI scribe technology was not widely used – but rapid adoption in primary care may change how clinicians record their notes. The dataset could also be used to compare GP-authored notes with those generated by ambient AI scribes.

This study created a reusable archive of telephone consultations with permission for secondary analysis, subject to further ethical approval. This resource may reduce the burden on practices and patients for new data collection and may lower the costs of future studies.

## AI Statement

Generative AI (ChatGPT 4o, 4o1, 4.5, 5) was used in the editing process of the manuscript and amending Stata code. PJE takes overall responsibility for checking any AI outputs were accurate and not plagiarised.

## Ethical approval

NHS research ethical approval was obtained reference number: 22/SW/0139

## Supporting information

Supplementary healthcare professional questionnaire

Supplementary patient questionnaire

## Data Availability

Telesafe is a controlled dataset as participants are potentially identifiable by their voices. For more information on accessing the dataset see https://www.bristol.ac.uk/primaryhealthcare/researchthemes/telesafe/

## Acknowledgements

The authors thank all the participating practices, clinicians and patients for their contribution towards this research.

## Supplementary Tables

**Table S1.** International Classification of Primary Care (ICPC-2) problem types in Telesafe and One in a Million. Data for Telesafe include all 96 usable consultation recordings. Data for One in a Million are as reported by Jepson et al.⁶ *Cons* = consultations. Note: this table uses ICPC-2 (rather than ICPC-3 used in the main text), as the One in a Million study was published before the release of ICPC-3.

| ICPC-2 Problem Category | Telesafe<br>(telephone cons.) |  |  | One in a Million<br>(face-to-face cons.) |  |  |
| --- | --- | --- | --- | --- | --- | --- |
|  | n | % of problems | Number of cons. with this problem type | n | % of problems | Number of cons. with this problem type |
| A - General and unspecified (A) and process codes (-) | 29 | 18.0% | 25 | 31 | 6% | 25 |
| B - Blood, blood forming organs and immune mechanism | 3 | 1.9% | 4 | 9 | 1.7% | 9 |
| D - Digestive | 15 | 9.3% | 12 | 53 | 10.2% | 52 |
| F - Eye | 2 | 1.2% | 2 | 6 | 1.2% | 6 |
| H - Ear | 2 | 1.2% | 2 | 11 | 2.1% | 11 |
| K - Circulatory | 18 | 11.2% | 18 | 39 | 7.5% | 37 |
| L - Musculoskeletal | 16 | 9.9% | 17 | 94 | 18.1% | 89 |
| N - Neurological | 6 | 3.7% | 6 | 22 | 4.2% | 21 |
| P - Psychological | 7 | 4.3% | 7 | 63 | 12.2% | 63 |
| R - Respiratory | 6 | 3.7% | 7 | 48 | 9.3% | 46 |
| S - Skin | 12 | 7.5% | 12 | 45 | 8.7% | 42 |
| T - Endocrine/metabolic and nutritional | 14 | 8.7% | 14 | 22 | 4.2% | 22 |
| U - Urinary | 9 | 5.6% | 9 | 23 | 4.4% | 23 |
| W - Pregnancy, childbearing, family planning | 4 | 2.5% | 4 | 21 | 4.1% | 21 |
| X - Female genital system | 12 | 7.5% | 12 | 20 | 3.9% | 20 |
| Y - Male genital system | 5 | 3.1% | 4 | 11 | 2.1% | 10 |
| Z - Social problems | 1 | 0.6% | 1 | 0 | 0% | 0 |
| **Total Problems** | **166** |  |  | **518** |  |  |
| **Total Consultations** | **96** |  |  | **327** |  |  |

**Patient questionnaire [separate file]**

**Healthcare professional questionnaire [separate file]**

