## Supplementary healthcare professional questionnaire for "The Telesafe archive: creating a database of UK primary care telephone consultations"

After an ethics amendment the study's name was updated to Telesafe

University of BRISTOL

| Study team use | HCP ID |
|---|---|
| Date received by study team:<br>Date logged:<br>Date data entered: | |

ONE IN A MILLION TELECON

### Healthcare professional Survey

Today's Date: [ ]

#### 1. Your status/role

| | | | |
|---|---|---|---|
| A partner in your surgery | ☐ | Pharmacist | ☐ |
| A salaried GP | ☐ | Paramedic | ☐ |
| Practice nurse | ☐ | Physicians Associate | ☐ |
| Nurse practitioner | ☐ | Foundation Doctor | ☐ |
| GP Registrar | ☐ | Other, please give details below | ☐ |

#### 2. What year did you first work as a health care professional in any general practice?

[ ]

#### 3. How long have you worked in <u>this</u> practice?

________ years _________months

#### 4. About you:

| Male | Female | Other | Prefer not to say |
|---|---|---|---|
| ☐ | ☐ | ☐ | ☐ |

#### 5. What year were you born in?

[ ] year

#### 6. Your working hours:

[ ] half-day sessions per week

**7. What is your ethnicity?**

| | | | |
|---|---|---|---|
| Asian | ☐ | Mixed | ☐ |
| Black | ☐ | White | ☐ |
| Chinese | ☐ | Other | ☐ |
| Prefer not to say | ☐ | | |

**9. What is your main language?**

| | |
|---|---|
| English | ☐ |
| Other (please specify) | ☐ |

**10. I give VERBAL safety-netting advice in my consultations**

| Rarely (<25%) | Sometimes (25-50%) | Often (50-70%) | Most of the time (70-90%) | Almost always (>90%) |
|---|---|---|---|---|
| ☐ | ☐ | ☐ | ☐ | ☐ |

**11. I give WRITTEN safety-netting advice in my consultations**

| Rarely (<25%) | Sometimes (25-50%) | Often (50-70%) | Most of the time (70-90%) | Almost always (>90%) |
|---|---|---|---|---|
| ☐ | ☐ | ☐ | ☐ | ☐ |

**12. Safety-netting advice is an important part of my consultations and is overall beneficial to patient care**

| Strongly disagree | Disagree | Neither agree or disagree | Agree | Strongly agree |
|---|---|---|---|---|
| ☐ | ☐ | ☐ | ☐ | ☐ |

**14. Safety-netting advice overall leads to increased patient demand**

| Strongly disagree | Disagree | Neither agree or disagree | Agree | Strongly agree |
|---|---|---|---|---|
| ☐ | ☐ | ☐ | ☐ | ☐ |

**15. The main reason I give safety-netting advice is:**

| Patient care | Mostly patient care some medico-legal cover | Equal patient care and medico-legal cover | Mostly medico-legal cover some patient care | Medico-legal cover |
|---|---|---|---|---|
| ☐ | ☐ | ☐ | ☐ | ☐ |

**Thank you.**
