## Supplementary patient questionnaire for "The Telesafe archive: creating a database of UK primary care telephone consultations"

After an ethics amendment the study's name was updated to Telesafe

| Practice team use | Study team use |
|---|---|
| Date of telephone appointment:<br>Reference:<br>Practice ID:<br>HCP ID:<br>EMIS ID: | Study ID code<br><br>Date received<br>Date logged<br>Date data entered |

### Patient Questionnaire

Please complete this questionnaire thinking about your latest telephone consultation with a healthcare professional (e.g GP, nurse, paramedic, pharmacist) at your GP practice.

| |
|---|
| **Please write today's date:** |

**1. How long after initially trying to book an appointment did your telephone appointment take place?**

| | |
|---|---|
| The same day | ☐ |
| The next day | ☐ |
| A few days later | ☐ |
| A week or more later | ☐ |
| Can't remember | ☐ |

**2. What were the main things you wanted from your recent telephone appointment?** ***Please ✓ all that apply***

| | | | |
|---|---|---|---|
| To find out what is wrong or get or a diagnosis | ☐ | For a routine check on a long-term health condition | ☐ |
| For reassurance | ☐ | To have a review of my medicines | ☐ |

| | | | |
|---|---|---|---|
| To ask for tests or investigations | ☐ | To ask for a referral | ☐ |
| To get the results of tests/investigations | ☐ | To follow-up on a previous consultation? | ☐ |
| To get treatment (e.g prescriptions, procedures) | ☐ | I wanted a face-to-face consultation | ☐ |
| Something else/other (please give details) | | | |

3. **How long have you had the main condition / symptom / problem that you discussed in your telephone appointment?** *Please ✓ one box*

| | | | |
|---|---|---|---|
| 1 week or less | ☐ | 6 months or more | ☐ |
| More than 1 week but less than 1 month | ☐ | Not applicable | ☐ |
| One month or more, but less than 6 months | ☐ | Not sure/can't remember | ☐ |

4. **Is this the first time you have consulted a healthcare professional for this problem?**

| | | | |
|---|---|---|---|
| yes | ☐ | No | ☐ |

5. **Thinking about appointments with healthcare professionals <u>in general</u>, do you have a preference for the type of appointment?**

| | | | |
|---|---|---|---|
| I prefer a telephone consultation | ☐ | I prefer a home visit | ☐ |
| I prefer a video call consultation | ☐ | I have no preference | ☐ |
| I prefer a face-to-face consultation at the surgery | ☐ | I don't know | ☐ |
| Other (please give details) | | | |

6. **Thinking about <u>this particular</u> telephone appointment with your healthcare professional, were you happy to have a telephone appointment on this occasion?**

| | | | |
|---|---|---|---|
| I was happy with a telephone appointment | ☐ | I would have preferred a home visit | ☐ |
| I would have preferred a video call | ☐ | I had no preference | ☐ |
| I would have preferred a face-to-face appointment at the surgery | ☐ | I don't know | ☐ |
| Other (please give details) | | | |

7. **My telephone appointment was with:**

| | | | |
|---|---|---|---|
| A GP | ☐ | A nurse | ☐ |
| A paramedic | ☐ | A pharmacist | ☐ |
| Someone else (please give details below) | ☐ | I don't know/not sure | ☐ |

8. **What happened in your telephone appointment?**

| | *Please ✓ one box to show the strength of your agreement with each statement* | **Strongly agree** (1) | **Agree** (2) | **Neither agree nor disagree** (3) | **Disagree** (4) | **Strongly disagree** (5) |
|---|---|---|---|---|---|---|
| **The healthcare professional I spoke to:** | | | | | | |
| a | Was helpful | | | | | |
| b | Was respectful and treated me with dignity | | | | | |
| c | Was knowledgeable about and understood my health condition/problem | | | | | |
| d | Was clear and easy to understand | | | | | |
| **I was given a full explanation, in clear language about:** | | | | | | |
| e | What caused my condition/problem | | | | | |
| f | The benefits and possible disadvantages of treatment | | | | | |
| **I was given the opportunity to:** | | | | | | |
| g | Discuss the problems in my life | | | | | |
| **I was given:** | | | | | | |
| h | Reassurance about my condition | | | | | |
| i | Advice about my health / condition | | | | | |

| | **What happened? Please tick all that apply**<br>*Please ✓ one box to agree or disagree with each statement* | **Yes** (1) | **No** (0) |
|---|---|---|---|
| j | A face to face consultation was arranged or agreed | | |
| k | Tests or investigations were arranged or agreed | | |
| l | I was given a results of test/investigations | | |
| m | I was offered advice about managing my condition / symptoms / pain | | |
| n | I had a check-up on my health condition(s) | | |
| o | I was given a prescription or other treatment for my condition or symptoms | | |
| p | I was given a referral to another doctor / specialist / therapist | | |

9. **Did the healthcare professional say what to do if your problem did not improve or got worse?**

| | | | |
|---|---|---|---|
| Yes | ☐ | Not sure | ☐ |

| | | | |
|---|---|---|---|
| No | ☐ | Does not apply | ☐ |
| If yes, what did they say? | | | |

**10. Did the healthcare professional prescribe any medicines (or give you a prescription for any medicines) as a result of this telephone appointment?**

| | | | |
|---|---|---|---|
| Yes (if yes please complete 10.a) | ☐ | No (if no please go to Q 11) | ☐ |

**10.a Did you collect these medicines?**

| | | | |
|---|---|---|---|
| Yes, all of them | ☐ | No, I decided not to take them | ☐ |
| Yes, some of them | ☐ | No, because I already had some | ☐ |
| No, I borrowed some from a friend/family member | ☐ | | |

**10.b Did you take these medicines?**

| | | | |
|---|---|---|---|
| Yes, all of them | ☐ | No, I decided not to take them | ☐ |
| Some of them | ☐ | No, I forgot to take them | ☐ |

**11. As a result of your recent telephone appointment, do you feel you are…**

| *Please ✓ one box to show the strength of your agreement with each statement* | **Much better** | **Better** | **Same** | **Less** | **Not applicable** |
|---|---|---|---|---|---|
| Able to understand your illness | | | | | |
| Able to cope with your illness | | | | | |
| Able to keep yourself healthy | | | | | |
| Able to cope with life | | | | | |
| | **Much more** | **More** | **Same** | **Less** | **Not applicable** |
| Confident about your health | | | | | |
| Able to help yourself | | | | | |

12. **How satisfied were you with this telephone appointment? (please try to answer this question thinking about *this* telephone appointment in particular)**
*Please tick (✓) one response*

| | | | |
|---|---|---|---|
| Very satisfied | ☐ | Dissatisfied | ☐ |
| Satisfied | ☐ | Very dissatisfied | ☐ |
| Neither satisfied nor dissatisfied | ☐ | | |

13. **Thinking about the healthcare professional you spoke to in your most recent telephone appointment, how well do you feel you know them**?

| | | | |
|---|---|---|---|
| Yes, very well | ☐ | No, not at all | ☐ |
| Yes, fairly well | ☐ | Don't know / can't say | ☐ |

14. **During your telephone appointment, did you have confidence and trust in the healthcare professional you spoke to?**

| | | | |
|---|---|---|---|
| Yes, definitely | ☐ | No, not at all | ☐ |
| Yes, to some extent | ☐ | Don't know / can't say | ☐ |

**<u>About you</u>**

*We would like to find out more about you. This is so that we can understand how experiences vary for different people.*

15. **Are you:**

| | | | |
|---|---|---|---|
| Male | ☐ | Other | ☐ |
| Female | ☐ | Prefer not to say | ☐ |

16. **How old are you?**

| |
|---|
| Please write in whole years: |

**17. Which of these qualifications do you have?** *(Please ✓ all the boxes that apply)*

| | | | |
|---|---|---|---|
| O levels / CSEs / GCSEs or equivalent | ☐ | Professional qualifications (for example teaching, nursing, accountancy) | ☐ |
| Apprenticeship | ☐ | Other vocational / work-related qualifications | ☐ |
| A levels / AS levels / Advanced Diploma or equivalent | ☐ | Overseas qualifications | ☐ |
| Degree / Higher degree / Higher Diploma or equivalent | ☐ | No qualifications | ☐ |

**18. Are you currently in paid work?**

| | | | |
|---|---|---|---|
| Employed / self-employed | ☐ | Looking after home or family | ☐ |
| Retired | ☐ | Unable to work due to illness / condition | ☐ |
| In full-time education | ☐ | Unemployed | ☐ |
| Prefer not to say | ☐ | | |

**19. What is your ethnicity group?**

| | | | |
|---|---|---|---|
| Asian | ☐ | Mixed | ☐ |
| Black | ☐ | White | ☐ |
| Chinese | ☐ | Other | ☐ |
| Prefer not to say | ☐ | | |

**20. What is your main language?**

| | |
|---|---|
| English | ☐ |
| Other (please specify) | ☐ |

**Thank you**

University of BRISTOL
